# Computer vision detects inflammatory arthritis in standardized smartphone photographs in an Indian patient cohort

**DOI:** 10.1101/2023.08.20.23294349

**Authors:** Sanat Phatak, Somashree Chakraborty, Pranay Goel

## Abstract

**Introduction:** Computer vision extracts meaning from pixelated images and holds promise in automating clinical tasks. Convolutional neural networks (CNN), deep learning networks used therein, have shown promise in X-ray images as well as joint photographs. We studied the performance of a CNN on standardized smartphone photographs in detecting inflammation in three hand joints.

**Methods:** We enrolled consecutive patients with inflammatory arthritis of less than two years duration and excluded those with deformities. Each patient was examined by a rheumatologist and the presence of synovitis in each joint was recorded. Hand photographs were taken in a standardized manner and anonymized. Images were cropped to include joints of interest. A reNrt-101 backbone modified for two class outputs (inflamed or not) was used for training. We also tested a hue augmented dataset. We report accuracy, sensitivity and specificity for three joints: wrist, index finger proximal interphalangeal (IFPIP), middle finger interphalangeal (MFPIP).

**Results:** The cohort had a mean age of 49.7 years; most had rheumatoid arthritis(n=68). The wrist (62.5%), MFPIP (47%) and IFPIP (41.5%) were the three most commonly inflamed joints. The CNN achieved the highest accuracy in being able to detect synovitis in the MFPIP (83%) followed by the IFPIP (74%) and the wrist (65%).

**Discussion:** We show that computer vision was able to detect inflammation in three joints of the hand with reasonable accuracy on standardized photographs despite a small dataset. Feature engineering was not required, and the CNN worked despite a diversity in clinical diagnosis. Larger datasets are likely to improve accuracy and help explain the basis of classification. These data suggest a potential use of computer vision in screening and follow-up of inflammatory arthritis.

## Introduction

Artificial intelligence is rapidly changing the landscape of healthcare in general and offers promising application in automating screening and follow-up of chronic disease.[1] Deep learning methods encapsulate the use of artificially created neural networks to learn functions that describe them. [2] Specifically, computer vision focuses on the extraction of meaning from pixelated images and videos. A convolutional neural network (CNN) is a type of deep learning neural network designed specifically for computer vision tasks. Using the mathematical operation called convolution allows a CNN to extract image features and consequently, for image recognition, classification, and segmentation. [3] Transfer learning is a technique that adapts a network which has previously been trained on a large dataset to effectively train that network on the images at hand.

CNNs are relatively recent but offer potential in automating various clinical tasks in healthcare. Most commonly, CNNs have been used to classify radiological images into different patterns (image classification), that allows faster and more accurate diagnoses.[4] A CNN (CheXNet) successfully detected pneumonias on chest radiographs.[5] In addition, CNNS are useful in segmenting medical images, helping doctors identify important features such as malignant tumors and inflammatory lesions. Clinical images are also valuable; a CNN to help diagnose diabetic retinopathy recently received Food and Drug administration (FDA) approval. [6] As computer vision techniques become more refined, each field of medicine and healthcare is likely to benefit.

The application of computer vision in inflammatory arthritis is nascent and has mainly focused on predicting erosive arthritis on X-rays, taking strides in automating scoring used in clinical trials.[7] Inflammatory arthritis presents with joint pain and swelling, and is typically diagnosed with history and examination during a visit to a physician. Detecting and recording the number of swollen joints is included in most classification criteria and disease outcome measures in these diseases. [8,9].

Most joints with synovitis do have visible signs of inflammation, including redness and swelling. We believed that these features would be amenable to detection by computer vision on standardized mobile-phone photographs of joint areas. A recently published CNN could distinguish inflammation in the proximal interphalangeal (PIP) by recognising the obliteration of creases on the joint. [10] Our ongoing study evaluates standardization methods for photography as well as the accuracy of computer vision in identifying inflamed joints. Subject to further validation, such approaches could be valuable digital incorporations into screening programs in communities with scarce access to rheumatology specialists, in addition to longitudinal follow-ups studies and clinical trials. [11]

We present pilot data on the performance of a CNN in detecting synovitis in a few selected joints in our ongoing study. We expanded the analysis to three joints: the wrist, 2nd/ Index finger proximal interphalangeal (IFPIP) and 3rd/ middle finger PIP (MFPIP), selected based on a relatively higher prevalence of involvement in the dataset. We evaluated the performance of this CNN without incorporating feature engineering.

## Methods

### Patient selection

We enrolled patients at the rheumatology outpatient clinic at the KEM Hospital, Pune as well as from a private rheumatology practice in Pune, India. We included consecutive patients with inflammatory arthritis involving the hand joints, of less than two years duration. Long standing arthritis was excluded in this dataset to prevent confounding by joint deformity. Permissible clinical diagnoses included but were not limited to rheumatoid arthritis (RA), psoriatic arthritis(PsA), systemic lupus erythematosus (SLE), Sjogren syndrome (SS) and chronic viral arthritis. We excluded patients with visually appreciable deformities, co-existing nodal osteoarthritis, and systemic sclerosis.

Demographic details recorded included age, gender; clinical details included duration of disease; clinical diagnosis; extra-articular features especially co-existent skin involvement. The patient’s and doctor’s global perception at that clinical visit (on a visual analogue scale) and results of markers of inflammation (ESR, CRP or both) were recorded. An independent clinical examination was conducted by a trained rheumatologist. The presence of synovitis was recorded as binary (Yes/No) opinion, based on inspection, palpation and range of motion testing in each of 15 joints in each hand (four distal interphalangeal, four proximal interphalangeal, one interphalangeal of thumb, five metacarpophalangeal and one wrist). For controls, only demographic information including age and sex was recorded (not used in this analysis).

### Hand photographs acquisition and standardization

Photographs of both hands from the dorsal and palmar aspect were taken by a trained research staff in standardized conditions. We modified a foldable photo studio light box used for product photography of dimensions 26 x 26 x 26 cm with an inbuilt LED light system and white background to ensure uniform camera placement on the top. The modified studio box was kept on a desk and patients sat in a chair to ensure appropriate placement of the hand. All photographs were taken using an iPhone 11 (Apple Inc, California, USA).

#### Data entry and anonymization

A unique identity generator was used to create randomly generated identifiers. [12] Two thousand unique IDs have been generated for patients with arthritis and controls each. Two IDs were generated for each subject: a personal ID (IDP), linked to personal information of the patient, and a study ID (IDS), linked to photographs and clinical information. The key to linking these two is securely stored with SP. Photographs were labeled with the IDS and suffixed as per hand laterality and view. Patient identifying data was entered with the IDP and clinical data related to synovitis prevalence with IDS. Only the anonymized, labeled photographs and non-identifying information was shared.

#### Joint detection and image cropping

**[Figure 1]**We used Media pipe, an open source library for tracking the key points of hands.[13] Media pipe can be used to locate joints of interest in clinical images of the hands. (x,y) coordinates of joints were extracted and used to isolate cropped images of the joints of the index finger proximal interphalangeal joints (PIP) and middle finger proximal interphalangeal joints (MFPIP), and the wrists.

**Figure 1:**
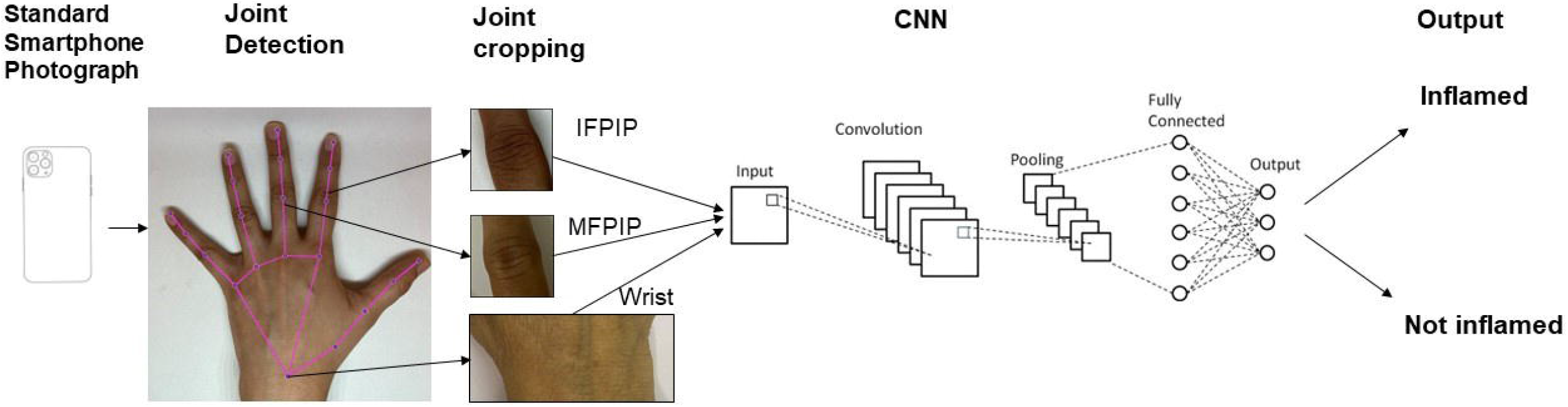
Schema of photograph processing and outputs from the convolutional neural network

#### Neural networks

The main model used for training was a ReNet-101 backbone modified for two class outputs: “Inflamed” or “not”. Transfer learning was used throughout. The dataset was split either 80:20 or 70:30 between training and validation sets. Typically, hyperparameters were varied during each training run: Batch sizes ranged from 32 to 64, training epochs numbered less than 100, learning rate was on the order of 1e-4, solvers were either “adam” or “sgdm” and early stopping was used when reasonable. Image augmentation typically included a random rotation up to 20 degrees both clockwise and anticlockwise, a random scaling between 0.8 and 1.2, random reflection about the X axis and a random X- or Y-shift up to 10 pixels. Neural networks were developed using the Deep Learning Toolbox in MATLAB. (MATLAB version: R2023b, Natick, Massachusetts: The MathWorks Inc.; 2023.)

#### Hue adjustment for skin tone

It is widely known that human skin varies in hue roughly between 6 and 34 degrees. (Skin Color Analysis. University of Edinburgh. 2001) A hue-augmented dataset was created as follows: The RGB image was converted to HSV format and a random hue component in the range 6 and 34 degrees was applied: (0.1 - 0.016) * (rand - 0.5); the modified image was clipped for hue values lying outside (0.16, 0.1). Each image was augmented for hue change once. [Figure 2]

**Figure 2:**
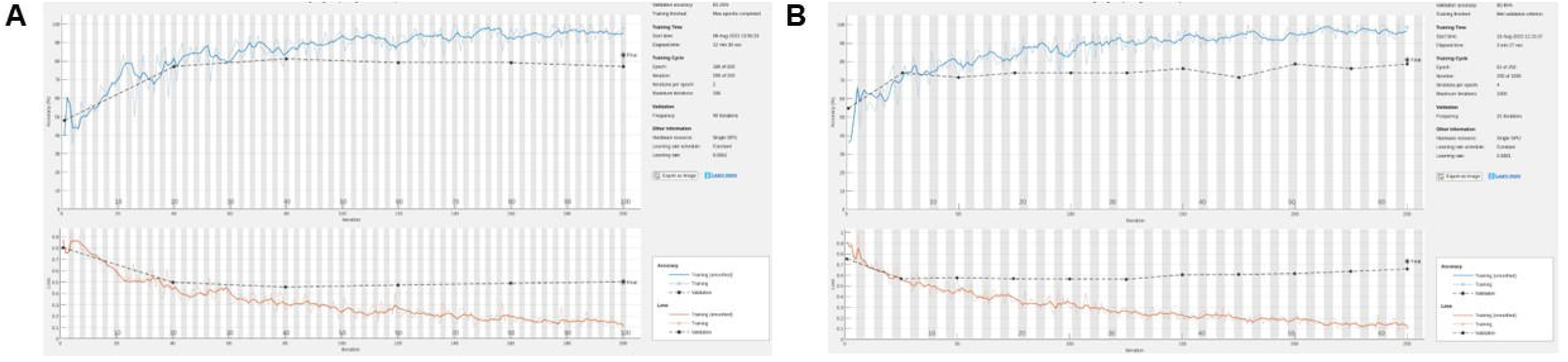
Hue augmentation for skin tone: Representative hue-augmented pairs with random hue variation between 6 and 34 degrees. A: index finger Proximal interphalangeal joints, B: Middle finger interphalangeal joints, C: Wrist joints. Note the henna on some images which did not change results.

Statistics: Patient data, especially the distribution of synovitis are presented as frequencies, and mean (standard deviation) for continuous variables. We report accuracy, sensitivity and specificity of the CNN in detecting synovitis considering rheumatologist opinion as gold standard.

#### Ethics

This study received ethics permission from the KEM Hospital Research Centre ethics committee (KEMHRCEC/2018) and a waiver from the IISER Ethics Committee for Human Research. A Data Sharing Agreement has been signed between the institutions for this study. All patients signed an informed consent document with special permission for storage of photographs in the photo repository for 30 years. This study has been registered with the Clinical Trials Registry of India (CTRI/2020/08/027129).

## Results

### Patient characteristics

We included 100 consecutive patients seen at less than two years of onset of arthritis. Mean age was 49.7 years, ranging from 18 to 76 years. Mean Erythrocyte sedimentation rate (ESR) was 42.8 (28.2 mm Hg) and C-reactive protein was 18.4 mg/dL. Sixty-eight had RA, eight had peripheral spondylarthritis, six had PsA, 13 had connective tissue diseases (three with SLE, four with SS, six with undifferentiated or mixed connective tissue disease). Five patients had chronic post-viral arthritis.

Nineteen patients had synovitis in only one hand, while 81 had bilateral disease. Of 200 hands studied, 52 had one joint swollen, 33 had two joints, 38 had three while 58 had polyarthritis (four or more). The wrist was the most common joint to be swollen (62.5% hands) followed by the MFPIP (47%) and IFPIP (41.5%). DIP joint swellings were relatively rarer. [Table 1]. All patients were able to lay their hands flat inside the photo box, since deformities were excluded. Since these were all adult participants and the depth of the box was constant, the aspect ratio was not altered and images were distortion free; thus no editing was required.

**Table 1:** Patient characteristics- Distribution of synovitis in the dataset (n=100)

| Swollen joints | Left hand<br>(n=100) | Right hand<br>(n= 100) | Total<br>(n= 200) |
| --- | --- | --- | --- |
| Wrist | 61 | 64 | 125 (62.5%) |
| MCP 1 | 5 | 15 | 20 (10%) |
| MCP 2 | 14 | 23 | 37 (18.5%) |
| MCP 3 | 16 | 21 | 37 (18.5%) |
| MCP 4 | 5 | 3 | 8 (4%) |
| MCP 5 | 3 | 4 | 7 (3.5%) |
| IP 1 | 8 | 19 | 27 (13.5%) |
| PIP 2 (IFPIP) | 41 | 42 | 83 (41.5%) |
| PIP 3 (MFPIP) | 44 | 50 | 94 (47%) |
| PIP 4 | 17 | 21 | 39 (19.5%) |
| PIP 5 | 17 | 20 | 37 (18.5%) |
| DIP 2 | 1 | 2 | 3 (1.5%) |
| DIP 3 | 2 | 1 | 3 (1.5%) |
| DIP 4 | 2 | 0 | 2 (1%) |
| DIP 5 | 1 | 1 | 2 (1%) |
| No swollen joint | 9 | 10 | 19 (9.5%) |

IP: Interphalangeal; PIP: Proximal interphalangeal; DIP: Distal interphalangeal; MCP: metacarpophalangeal

### CNN performance

The CNN achieved the highest accuracy with the MFPIP (83%), followed by the IFPIP.[Table 2] Accuracy in detecting wrist synovitis was lower in comparison. Note that the percentages in the table are largely representative. Given that the numbers of images in the dataset are relatively small for deep learning, particular training runs give rise to somewhat different accuracies, typically varying about 5% - depending on the particular samples being trained. We found a sensitivity and specificity of the CNN in detecting synovitis, taking the rheumatologist’s opinion as gold standard. The specificity was the lowest at the wrist joint. [Figure 3]

**Table 2:**
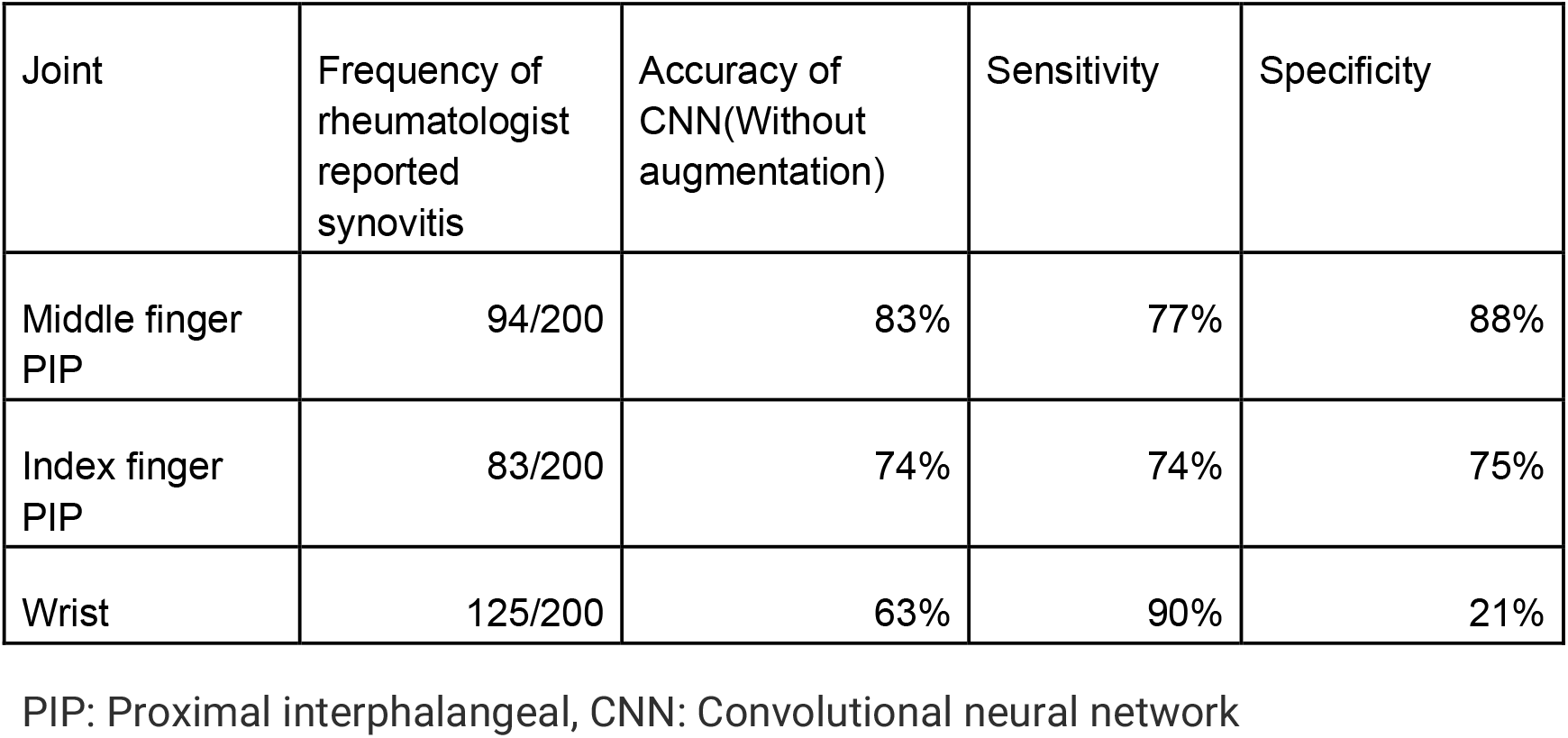
Performance of convolutional neural network(CNN) in detecting synovitis in three commonly involved joints.

**Figure 3:**
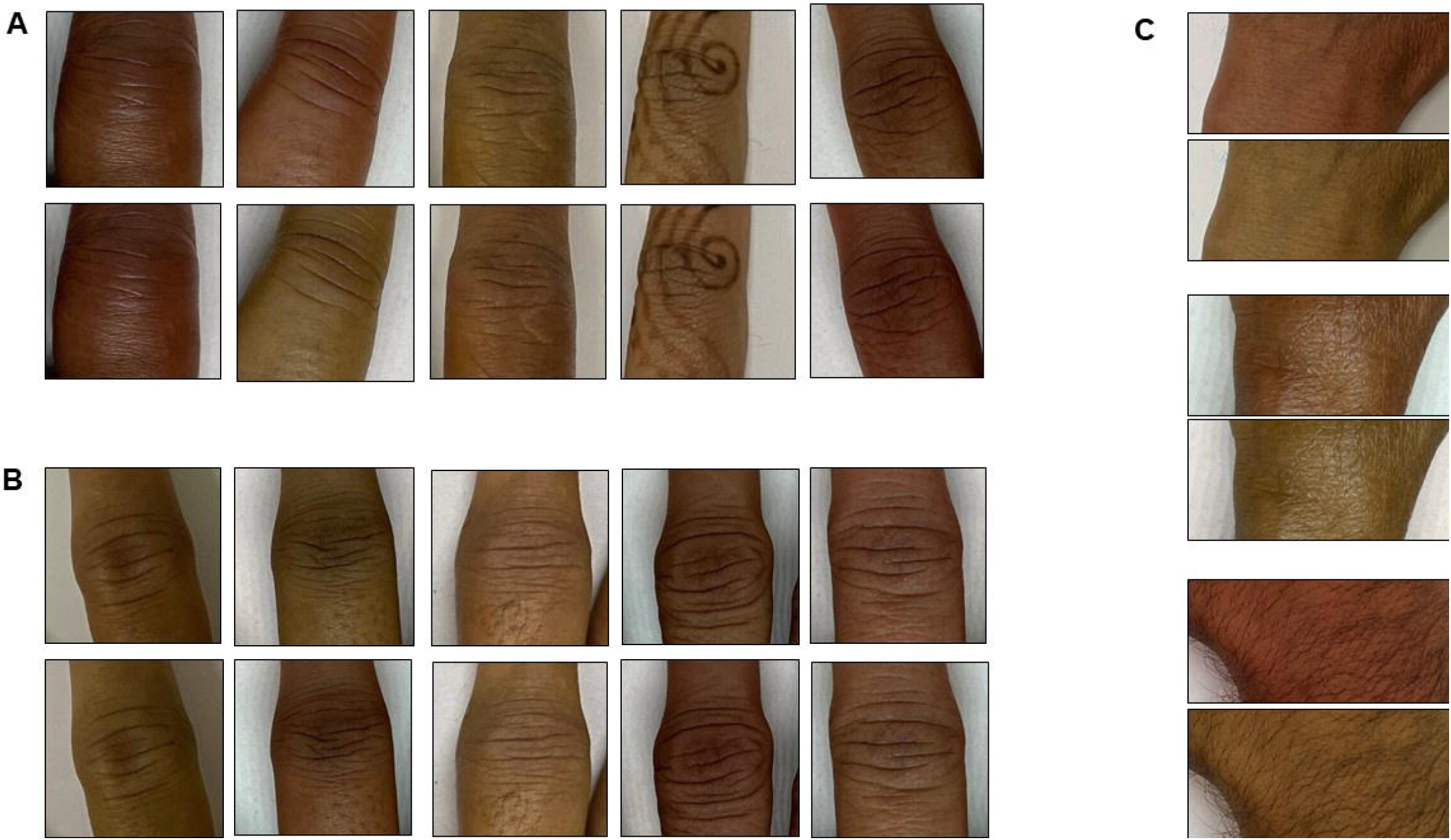
A-Representative training graph for naive (no hue augmentation) Middle finger PIP. On this run, accuracy=83%, sensitivity=77% and specificity=88%. B: representative training graph for naive (with hue augmentation) MFPIP. On this run, accuracy=81%, sensitivity=65% and specificity=92%

We created a dataset in a skin tone augmentation was applied in a 1:1 ratio, that is, each original image was augmented with one image with a modified hue (see Methods). Training was then carried out on this data. The rationale was to test the training by augmentation the data with different samples of skin tone. This resulted in a substantially higher validation accuracy (92-95%), especially if a larger ratio of original to hue-modified images was used. However, it is not clear if those results are artefacts of having too many modified images common between training and validation sets, even though a conservative 1:1 ratio of hue augmentation was used. We therefore retrained the networks with training set containing hue augmented images, while the validation set contained only the original images. With this training protocol the results were comparable to the case above (without hue augmentation).

## Discussion

In this proof-of-concept analysis, we demonstrated that computer vision was able to distinguish inflammatory arthritis, with reasonable accuracy in three selected synovial joints of the hand on standardized photographs. The CNN worked despite in a real-world setting despite the heterogeneity in the etiology of inflammation, and without feature engineering. Accuracy remained constant using skin tone augmentation using modified hue. These data increase confidence in the utility of CNNs across disease and ethnic groups. We provide credibility to the potential future use of applying computer vision on smartphone photographs as a screening and follow-up tool in inflammatory arthritis.

CNNs have been used previously in rheumatology, mainly in automating analysis of microscopic immunofluorescence images as well as for radiographic images, achieving agreement close to human expert readers.[14] A ‘cascade’ of CNNs could discriminate healthy joints with an accuracy comparable to trained rheumatologists from hand X-ray images [15] Similarly, CNNs have been trained to perform scores such as the modified van der Heijde Sharp Score for joint erosions on hand radiographs. [16] A large body of work using both MRI and X-ray images in detecting and scoring rheumatoid arthritis erosions with various learning mechanisms and classification techniques is now available. [7] While these are highly useful, most are use cases for specialists once a diagnosis is made and require expensive imaging technologies. In more resource limited settings such as ours, there is a felt need for applications at the screening and diagnosis level. Preassessment using tele-health for efficient referral can help optimize time for rheumatologists. [18] Smartphone based photographs can be easily taken even before the patient has access to imaging technologies, and at negligible cost. Promising steps have been taken in establishing the utility of smartphone camera sensors in assessment of physical function in RA. The PARADE study demonstrated their utility in capturing functional ability in wrist movements and gait in patients [17]. The innovative TELERA randomized trial is evaluating the possibility of using app-based outcome measures in remotely following up patients with arthritis. [19]

Recently, a CNN that recognized joint swelling was shown to be efficacious as a digital biomarker. [10] The authors used cropped-out images of the PIP joints in a similar sized dataset as ours. The patient population in the study by Hugle et al was more homogenous, with all patients satisfying criteria for RA although joint swelling distribution; ethnic and skin colour characteristics of the cohort were not commented upon but was presumably Caucasian. Our entire patient group was South Asian, all falling in grade 3-4 on the Fitzpatrick classification. Our results expand on this study by showing a preservation of CNN accuracy not only on additional joints (IFPIP and wrist) but also across diseases and ethnic groups, especially in darker skin where inflammatory erythema may not be as readily visible. Our photos included those with henna markings and hand jewelry, which did not reduce accuracy.

The CNN performance did not improve significantly by training on the hue augmented images. Thus we reason that while hue augmentation is not an effective strategy for training, however, the results of training (that is, the trained networks) will be effective for applying across populations with varying skin tones. Skin hue modification and its implications for training and prediction is likely to be an interesting area to pursue in future work, especially to improve generalisability across populations.

Like in any black box AI technology, we are not certain what parameters were used to classify synovitis by the CNN, and one can only speculate that a combination of color, shape and contour changes was responsible. Hugle et al included identification and extraction of dorsal finger folds from the joint images. [10] Intuitively, this methodology may not extrapolate to synovial joints that do not have naturally occurring skin folds, such as MCP, wrists, knees or ankles. Our methodology hopes that any relevant features will be learned in the training. For example, it has learnt to ‘overcome’ confusion due to henna and rings. Even without feature engineering, our CNN achieved similar accuracy to their dataset, supporting a more general approach. The joint shape may be important since wrist inflammation was picked up less accurately in our analysis. Larger, uniform datasets would be required for a deeper delve into trying to explain the basis for the classification.

Despite being an initial analysis of an ongoing collection, our study has certain strengths: our patient distribution is heterogeneous and reflects a real-world rheumatology practice. We used carefully standardized photographs using the same phone camera. Weaknesses include a small dataset. This precluded an analysis of the other joints of the hand since such a small fraction was swollen. Our gold standard of a rheumatologist’s assessment is also subjective; further work would include the evaluation of the CNN’s performance against more objective measures such as MRI detected synovitis. Finally, since we included only early arthritis, the performance in the presence of joint deformity or co-existing osteoarthritis, both important clinical confounders, cannot be commented upon at this stage. Nevertheless, our pilot results increase confidence that larger, good quality and diverse datasets would improve reliability of computer vision-based applications in inflammatory arthritis. These would also enable determining individual accuracies for different joints, as well as Bayesian methodologies to improve synovitis detection.

In conclusion, we show that computer vision could detect synovial joint inflammation with reasonable accuracy in standardized smartphone photographs of joint areas. With larger datasets, this technology has potential in being a valuable remote tool in screening and follow up for inflammatory arthritis.

## Author Contributions

SC: Data curation, processing images for joint identification and cropping. PG: Classification using deep learning. SP: Data collection, PG and SP: Writing the paper.

## Data availability statement

Anonymized and labelled photographs are stored with SP and would be available on request.

